# Proof-of-implementation of the SMART2D intervention: findings from a cluster randomised feasibility trial for prevention and control of type 2 diabetes in socio-economically disadvantaged neighbourhoods in Stockholm, Sweden

**DOI:** 10.64898/2026.09.30.26364359

**Authors:** Meena Daivadanam, Kristi Sidney Annerstedt, Helle Mölsted Alvesson, Linda Timm, Carl Johan Sundberg, Peter Delobelle, Josefien Van Olmen, Pilvikki Absetz, David Guwatudde

## Abstract

**Background:** Type 2 diabetes (T2D) disproportionately affects socioeconomically disadvantaged communities. Evidence on the feasibility and effectiveness of prevention and self-management strategies in complex contexts remains limited.

**Objective:** To report effectiveness outcomes from a feasibility trial comparing the SMART2D intervention with routine primary care for T2D prevention and control in two neighbourhoods in Stockholm.

**Methods:** A cluster-randomized feasibility trial with hybrid type 1 design was conducted in two neighbourhoods among individuals at high risk of T2D (n=84) or with diagnosed T2D (n=58), allocated to an integrated care arm (n=70; personal coaching and community outreach) or facility-only arm (n=72; routine care). Primary outcomes were glycaemic control (participants with T2D) and HbA1c reduction (participants at high risk of T2D); secondary outcomes were retention to care and diabetes incidence (participants at high risk of T2D). Analysis used intention-to-treat principles: difference-in-differences with mixed-effects models for HbA1c reduction, and modified Poisson regression (IRR, 95% CI) for other outcomes.

**Results:** Among participants with T2D, glycaemic control (adjusted IRR=0.92, 95% CI=0.89-0.95) and retention to care (adjusted IRR=0.91, 95% CI=0.91-0.92) were significantly lower in the integrated care arm. Among participants at high risk of T2D, integrated care showed a marginally significant, greater HbA1c reduction (adjusted 2.02, 0.20-3.84) and significantly higher retention to care (adjusted IRR=1.22, 1.15-1.29), plus fewer incident diabetes cases (0 vs. 2 in facility-only).

**Conclusions:** Integrated care (personal coaching, community outreach) benefited HbA1c reduction and retention among participants at high risk of T2D, whereas facility-only care improved glycaemic control and retention among participants with T2D.

## BACKGROUND

According to the International Diabetes Federation (IDF) Diabetes Atlas (2025), an estimated 11.1% of the adult population (20-79 years) globally is living with diabetes, with almost half remaining undiagnosed [1]. In Sweden, the setting for this study, diabetes prevalence is estimated at approximately 7.5% among adults with an age-adjusted comparative prevalence of diabetes around 5.8% and 28.5% of people with undiagnosed diabetes [1]. The majority of all reported diabetes numbers is attributed to type 2 diabetes (T2D) [1]. However, the prevalence varies by neighbourhood characteristics and has been found to be higher in socio-economically disadvantaged groups and suburbs [2–5]. There are no national prevalence estimates for populations at high risk for T2D based on representative samples or comparable methodology. The IDF Diabetes Atlas [2025] for example, does not give a single estimate, rather separate ones for Impaired Fasting Glucose (5%) and Impaired Glucose Tolerance (8%) to avoid double counting[6]. Moreover, the distribution of T2D risk in the population was found to be highly heterogeneous. Immigrant men aged 70–79 years, with low educational achievement and low income for example, had an 18% risk of developing T2D, compared with 0.5% risk among native women aged 40–49 years with high income and high educational achievement [7].

The population in Sweden like in other parts of Europe, has increasingly become multi-cultural and diverse and the diversity is complex, going beyond mere ethnic diversity [8]. These suburbs, particularly in the major cities like Stockholm are characterised by a complex interplay of multiple intersecting diversities seen in modern cities, described by Vertovec et al as *superdiversity* [9]. Originally defined for London, the concept has since been expanded to include similar cities, describing a dynamic interplay of variables affecting where, how, and with whom people live [10]. This includes new, small, and scattered immigrant groups of multiple national origins that are transnationally connected, socioeconomically differentiated, and legally stratified [10]. These patterns go well beyond earlier, simpler forms of ethnic diversity, such as a few large, stable, and clearly identifiable ethnic or national-origin communities [10]. In Sweden, foreign-born individuals defined as individuals who were either born abroad or having both parents born abroad account for about 19% or two million inhabitants in 2010 [10], increasing to 27% in 2024 [10]. Foreign-born residents in Sweden mainly come from Syria, Iraq, Finland, Poland, Iran, Afghanistan, Somalia, Yugoslavia, and those with a non-European background account for 57% of the total foreign-born population in 2024 [11].

The contextualisation and implementation of public health interventions become more complex in contexts of superdiversity, where the lack of a common language is only one of a myriad and complex range of challenges [12]. It is therefore unsurprising that the majority of the interventions focus either on Swedish-speaking populations or mono-ethnic groups such as foreign-born individuals from Somalia, Iran, Iraq or Syria [13,14]. Qualitative studies by this team and others have found that T2D patients experienced healthcare advice to be largely focused on clinical treatment [15,16]. Moreover, patients found it difficult to adapt food and physical activity advice to their own cultural context, while providers viewed this diversity in food, physical activity and treatment perceptions as challenging and perceived themselves as inadequately trained to handle it. [17]. However, ‘real world’ interventions should be applicable for ‘real world’ contexts with their complex population make-up and dynamics, not just parts of that population. While effective interventions are available for T2D prevention and management, little is known about their applicability for such superdiverse neighbourhoods and populations, in terms of how uniform a strategy or intervention should be; and how much adaptation is needed to address this superdiversity.

T2D care and prevention is dependent on the regular execution of self-management that includes different types of self-care and health behaviours [18]. Interventions to address self-management tend to be complex and multi-faceted, often requiring multi-actor or sectoral collaborations. This is challenging, particularly considering the need to test or explore the feasibility of ‘real world’ interventions over several cycles before they can become ‘standard’ care. Several reviews of T2D and prediabetes prevention trials have demonstrated their efficacy and effectiveness [19,20], while other reviews have focused on showing the effectiveness of T2D prevention in ‘real-world’ contexts in terms of effect size, scope of services, scalability potential, and long-term sustainability [21–23]. The real-world testing of interventions are usually seen to yield lower effect sizes than their testing under more controlled conditions [24,25], often attributed to two phenomena, namely *voltage drop* whereby interventions are expected to yield lower effects as they move from efficacy to effectiveness and into real world use; and *programme drift*, whereby deviations from written protocols occur during the real-world delivery of interventions leading to lower observed effects. This has been largely attributed to the dynamicity of the context adversely affecting the intervention and its implementation. Chambers et al. argue that understanding the changing context can help to continuously refine and improve interventions as they are sustained [26] Hence, feasibility trials have an important role in assessing the effectiveness and improving the sustainability of complex interventions. However, the time lag between trial implementation and evidence generation discourages the use of in-situ and multi-cycle feasibility tests which are often required to fine-tune interventions in ‘real world’ contexts [27,28].

The ‘Self-Management Approach and Reciprocal Learning for Type 2 Diabetes’ with acronym “SMART2D”, aimed to support self-management in T2D prevention and management in disadvantaged or under-resourced settings in South Africa, Uganda and Sweden [29–32]. The aim of the project was to develop an intervention with shared key functions and a good fit with the local context, needs, and resources of each of the three settings, subsequently tested in an adaptive implementation trial [29,31]. However, the challenges related to a superdiverse population and healthcare re-organisation in the Swedish setting, described later, necessitated a change from an implementation to a feasibility trial , including testing implementation and effectiveness in a limited sample in line with the recommendations made by Bowen et al [33].

This article describes the adaptations of the SMART2D multisite study protocol to a feasibility trial to assess proof-of-implementation and report the results thereof, in socio-economically disadvantaged and superdiverse neighbourhoods in Stockholm, Sweden.

## METHODS

### Study setting

This study was conducted in two socioeconomically disadvantaged neighbourhoods in Stockholm County, Sweden, with a significant immigrant population (at least 36%) [34]. These neighbourhoods included diverse nationalities, mainly from the Middle East and Africa. Residents had generally lower levels of educational backgrounds, low incomes, high unemployment rates, and poorer housing compared to other parts of Stockholm County [30,35,36]. The Care Need Index (CNI), indicating social deprivation, was also higher in these neighbourhoods compared to areas with average socioeconomic status in Stockholm County [37].

Standard diabetes care is provided by primary healthcare centres (PHCs) staffed by health professionals, including doctors and nurses, who refer patients to secondary and tertiary care services like dieticians, podiatrists, ophthalmologists, and endocrinologists when necessary [38]. Swedish diabetes prevention and treatment guidelines recommend regular eye exams, foot care, and monitoring of glycaemic control, blood pressure, and cholesterol to prevent and minimize diabetes-related complications [38]. Patients with T2D registered at a PHC receive subsidized care, with two annual follow-up visits: one with a doctor for tests like HbA1c, cholesterol, and blood pressure, and one with a diabetes nurse for lifestyle education [38]. While this is standard routine care for T2D, there are wide variations in diabetes care quality and outcomes between PHCs in Region Stockholm in terms of screening and follow-up practices (Harcke et al., 2022) and in patients’ odds of achieving risk-factor control, which are independently associated with socioeconomic status and country of birth [39]. Moreover, PHCs situated in socio-economically disadvantaged suburbs experience high staff turnover, limited resources, and additional challenges in managing ’difficult patients’, described by providers as persistently unable to achieve glycemic control or difficult to reach due to frequent travel or extended stays abroad [17].

### Study design

We used a hybrid Type 1 study to test the effectiveness of the SMART2D self-management intervention on T2D outcomes (primary aim) through a feasibility trial to assess its potential in a real-world setting with a superdiverse population (secondary aim) [40].

The feasibility trial was developed in line with the eight general areas of focus for feasibility studies outlined by Bowen et al. 2009 [33]. Findings on Acceptability, assessed using Sekhon’s Theoretical framework of Acceptability; Perceived demand, assessed through reach of screening; and Fidelity, assessed with a focus on interaction between the tele-health facilitators and participants; have been reported earlier by Timm et al 2022, 2020, and 2021, respectively [41–43]. The present study focuses on Limited effectiveness, while the remaining three, Adaptation, Integration and Expansion, have not been assessed.

We used the CONSORT 2010 statement: extension to randomized pilot and feasibility trials for reporting the results of our study [44].

### Randomization protocol

The trial design followed the multisite protocol outlined in Guwatudde et al (2018) and ISCRTN registry [31,45]. We conducted a cluster-randomized feasibility trial involving two collaborating PHC facilities. Each neighbourhood comprising a PHC facility and its surrounding catchment area were divided into two geographically distinct clusters, yielding four clusters in total. A cluster was defined as a PHC facility and contiguous postal codes roughly comprising half the population within a neighbourhood to facilitate local inspiration meetings. Within each neighbourhood, one cluster was randomized to receive the intervention (integrated care) and the other the control (facility-only) arm. A few overlapping postal codes between intervention and control clusters, where we assumed a high risk for contamination, were excluded. Figure 1 is a pictorial depiction of the clusters in the two neighbourhoods. As the society in Sweden is largely individualistic, the allocation assumed minimal spillover effect between integrated care and facility-only clusters in the same neighbourhood. Recruited participants residing in these areas were automatically allotted to the two arms based on their postal codes. Participants in the integrated care arms received facility strategies at their PHC as well as community based self-management support; while those in the facility-only arms received augmented usual care (described below).

**Figure 1:**
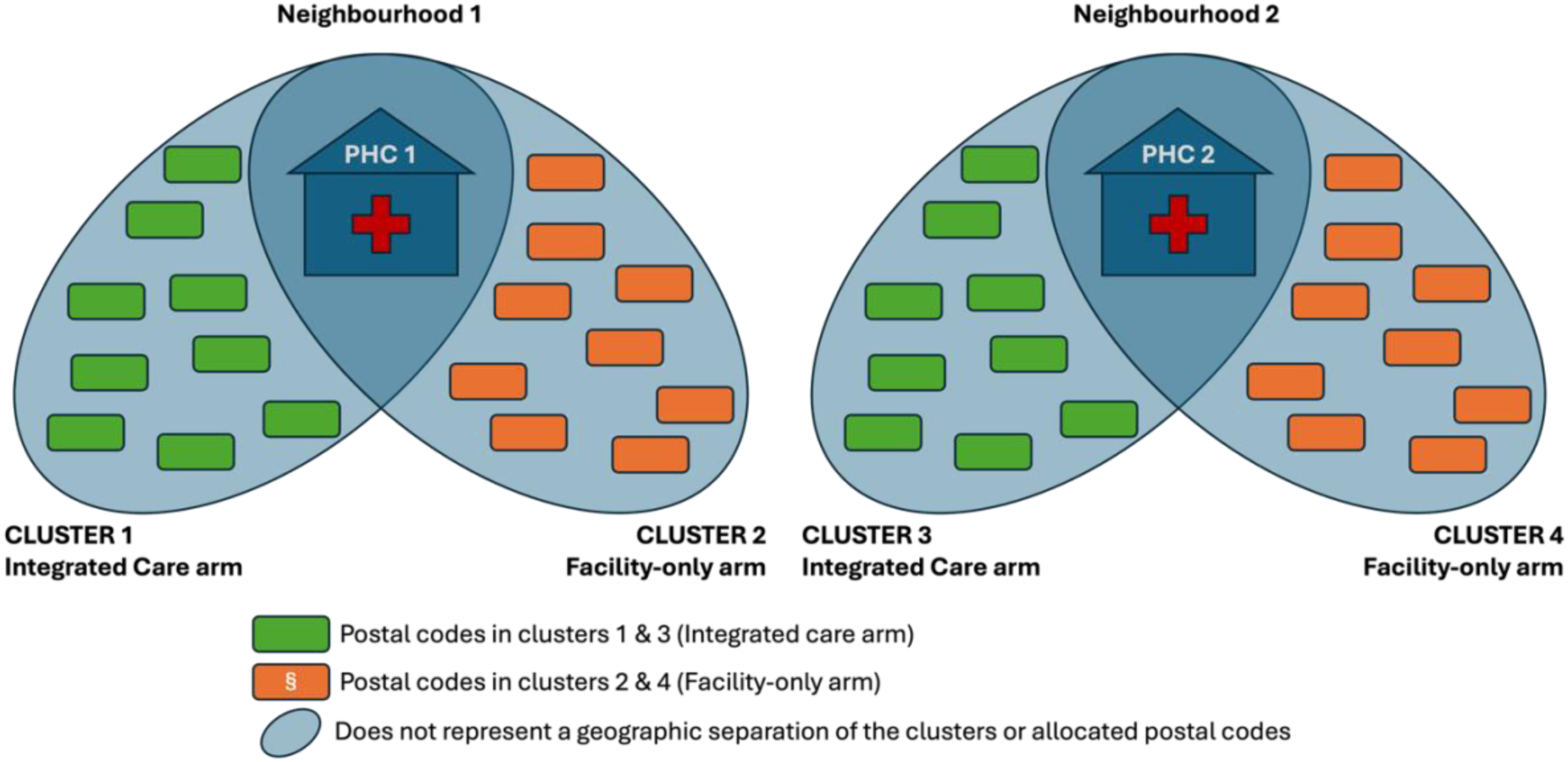
Visualisation of clusters in the feasibility trial

### Participant recruitment

Between March 2017 and June 2018, community screening was conducted [43]. During the preparation phase, the SMART2D team held planning meetings at participating PHC facilities, citizen service offices, and local organizations at the two study sites. Citizen service office employees helped mobilize individuals to the screening venues, where informational leaflets and brochures were distributed, designed in simple Swedish with illustrative pictures. Special flags and roll-up posters were displayed at the venues for visibility. Screening took place in shopping malls, citizens offices, premises of local organisations and ethnic cafes, women’s centres, and swimming facilities at varying times, including weekends. The screening involved administering a FINDRISC questionnaire [46] and a point-of-care HbA1c test. Scores ≥12/25 on the FINDRISC indicated a high risk for T2D. Those with HbA1c levels above 38 mmol/mol were referred to participating PHC facilities or advised visiting a PHC facility of their choice for further examinations. Those identified as high risk for T2D were invited to participate in the trial. Additionally, diabetes nurses from collaborating PHC facilities invited individuals with pre-diabetes or T2D registered at their PHC facility to participate in the study. Details about inclusion and exclusion criteria have been published elsewhere [31,45].

### The SMART2D intervention

The SMART2D intervention was based on a Theory of Change, which was outlined in the SMART2D self-management framework [30]. Facility strategies consisted of strengthening and standardizing existing or usual care as outlined in Guwatudde et al (2018), referred to as augmented usual care [31]. The self-management intervention consisted of three components assumed to provide the peer support function outlined in the trial protocol (Figure 2): 1) one-on-one telephonic health coaching delivered by trained facilitators via real-time audio calls delivered over the Skype platform, consisting of nine sessions focusing alternately on physical activity and diet; 2) selection of a person(s) by the participant to be involved and support activities discussed during the personal coaching sessions, designated as Care Companion(s) (CC); and 3) Inspiration meetings at community level together with the facilitators, nurses from the PHC facility, local authorities, outreach workers and invited experts on diabetes, nutrition and physical activity [29].

**Figure 2:**
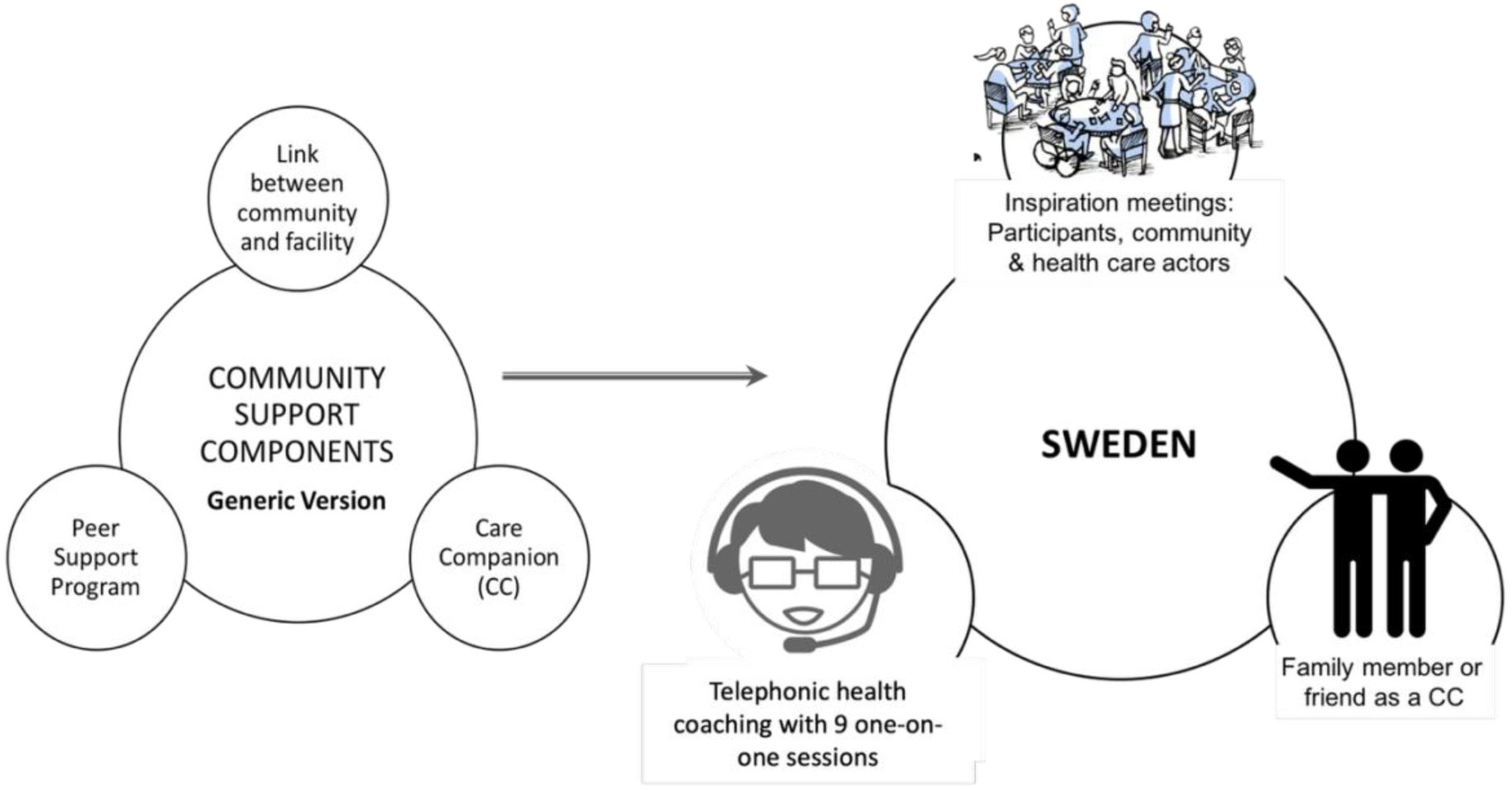
SMART2D intervention for Sweden in relation to the generic version of the intervention. Modified from the figure on Generic and contextualized community strategies and their components (Figure 1), Absetz et al. 2020 [47].

**Figure 3.**
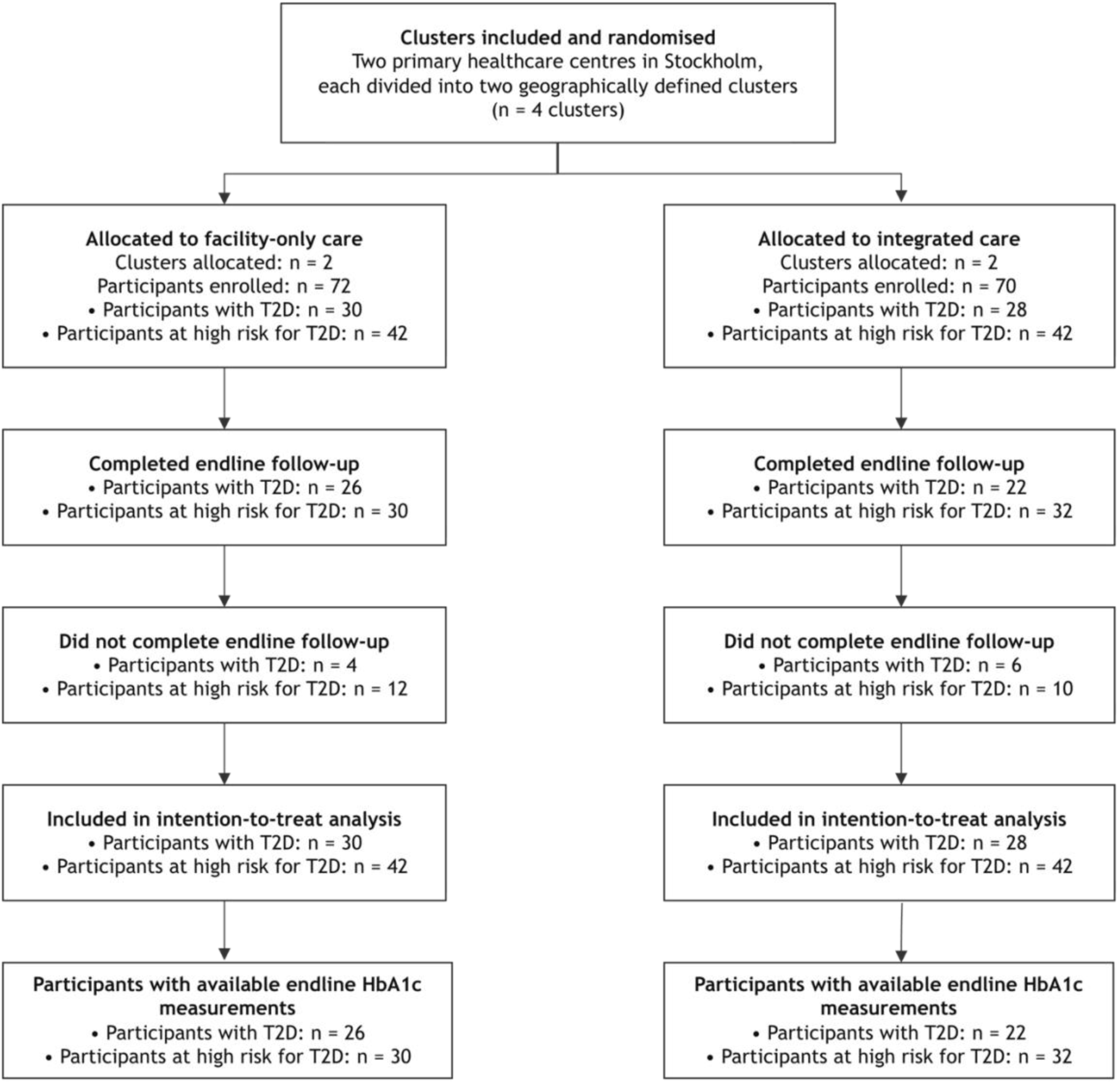
CONSORT flow diagram showing participants in the SMART2D Sweden feasibility trial.

### Telephonic Health Coaching

The telephonic health coaching was built on a strength-based approach using the brief motivational coaching format for behaviour change. A participatory, autonomy-supportive approach grounded in self-determination theory was used, aiming to identify existing healthy habits and self-management behaviour and help individuals to change behaviour [29,31]. A facilitator guide was used to deliver the structured telephonic coaching, consisting of an introductory and concluding session, together with seven alternating sessions focusing on physical activity or diet (Table 2). REDCap, an electronic data capture system was used to implement and manage sessions. All facilitators were trained and standard operational procedures were developed to ensure consistency in their administration of the sessions, which were conducted over the telephone using pre-paid Skype accounts. The sessions were planned at the convenience of participants, with some taking place during evenings or weekends. Each session lasted 10-45 minutes depending on the session and the needs of the participant. Some sessions were recorded for quality control. Each participant in the intervention group was assigned a contact person to facilitate the nine telephone sessions in different languages, including four facilitators for (easy) Swedish or English, one for Spanish and two facilitators for Somali and Arabic.

**Table 2.** Content of the Tele-Health Coaching Sessions

| Session # | Session content |
| --- | --- |
| 1 | Introductory session: Getting to know the program |
| 2 | Physical activity: Increase physical activity in daily life & reduce sedentary lifestyle |
| 3 | Diet: Healthy eating: Regular, balanced & healthy |
| 4 | Physical activity: Physical activity through the life-course |
| 5 | Diet: Fruit & vegetables: Improving regular consumption |
| 6 | Physical activity: Increasing your daily physical activity |
| 7 | Diet: Sugar: Reducing daily consumption |
| 8 | Physical activity: Finding a physical activity that suits you |
| 9 | Concluding session: Healthy lifestyle – moving forward |

### Care Companion

The role of the care companion (CC) was to provide emotional, practical, and ongoing support to individuals with T2D while visiting the health facility or participating in activities discussed in the tele-health coaching. CCs included friends, partners, spouses or children of the participant and could consist of different CCs for different activities. The companion also acted as a resource for the participants.

### Inspiration Meetings

In addition to the tele-health coaching sessions, the participants and their CC(s) were invited to two group meetings per locality organised as part of community outreach through the Community Link Team which structures on existing services in Sweden, namely, the primary care, citizen’s service offices run by the municipality and the research team [29]. At these meetings, participants could listen to or ask questions about T2D and lifestyle related issues to experts in the field. In addition, a diabetes nurse and dietician addressed questions about T2D and diet collected during previous tele-health coaching sessions. Participants were also provided information about facilities, activities and initiatives in the community organized by local civil society organisations or the municipality. The meetings also facilitated community linkages, with representatives from the citizens service offices run by the municipality and local PHC.

### Data collection and management

To ensure consistency and minimization of errors in data collection, standardized methods of data management were developed and used across the original SMART2D trial sites in Uganda, South Africa, and Sweden. The tools contained common elements for administration, i.e., the minimum set of common questions or data items to be collected at all country sites. In addition, each country site added site-specific questions of interest.

Once the content of data collection tools had been agreed upon and finalized, common data entry formats were developed in the REDCap data management system and pilot tested. Data fields or variable names and their formatting were maintained across sites to enable quick data merging at the end of the trial. Any required corrections were made before initiation of trial activities. As a standard operating procedure, data captured by tablets into REDCap were checked for completeness, inconsistencies, and any other errors at the end of each working day by a designated quality assurance (QA) manager. In case of errors, the responsible data collector attempted to correct, and/or contact the participant(s) where possible.

### Deviations from the original SMART2D protocol

Deviations from the original SMART2D protocol for implementation of the feasibility trial in Sweden are described in Table 3. The trial maintained fidelity to the main study outcomes as planned, but also focused on the implementation process using qualitative and mixed methods studies [41].

**Table 3.** Deviations to the original SMART2D study protocol

| Trial elements | As described in the original study protocol | Deviations as implemented | Rationale for deviations |
| --- | --- | --- | --- |
| Study participants | Diabetes & pre-diabetes | Diabetes & high-risk | Participants informed about their diabetes risk did not attend the PHC for confirmation of diagnosis (as high risk or prediabetes was not a diagnosis at the time) and there was a long lag-time between screening and start of intervention which increased those lost to follow-up. |
| Sample size | 245 in each arm | 58 in each arm | Reduction from five to two PHCs necessitated a reduction in sample size for limited effectiveness assessment. |
| Secondary outcome variables | ► Incidence of diabetes among participants with pre-diabetes;<br>► Incidence of adverse events, including hospitalisations due to hypoglycaemia- or hyperglycaemia, or complications of diabetes;<br>► Behavioural outcomes, including diet, physical activity, foot care, tobacco and alcohol consumption;<br>► Difference in out-of-pocket expenditure between baseline and end line and incremental (system level) cost of implementing the intervention;<br>► Among participants with diabetes, participant satisfaction with the diabetes treatment provided at the health facilities | ► Incidence of diabetes among participants with high-risk;<br>► Behavioural outcomes, including diet, physical activity, foot care, tobacco and alcohol consumption;<br>► Difference in out-of-pocket expenditure between baseline and end line and incremental (system level) cost of implementing the intervention;<br>► Among participants with diabetes, participant satisfaction with the diabetes treatment provided at the health facilities; | Modified to be relevant for the shorter intervention duration of 6 months. |
| Intervention design | Peer group programme | Peer group programme elements were converted to tele-health coaching but the basic content was maintained using the strength-based approach. | Based on piloting of the trial components |
| Duration of intervention | 1 year | 3 months | Loss of project time necessitated reduction in duration of intervention which aligned with the change to feasibility trial. |
| Criteria for completion of intervention | Participation in 1/3 of peer sessions [29] | Participation in minimum 3 sessions (either tele-health coaching sessions alone or in combination with community inspiration sessions) | Criteria modified to include participation in the community inspiration meetings which were around 2 hours duration and included interaction with the Community Link Team and experts. |

SMART2D commenced in 2014 with agreements to participate in the project from five primary PHC facilities in Sweden. However, by late 2015, a reform instituted by Region Stockholm saw a new administrative unit of mid-level managers being established who were also responsible for approval of research projects. The Swedish research team had to reapply to this new mid-layer administrative unit for approval of the trial and further reapply to the individual PHC facilities. This was further complicated by the human resource shortage in Stockholm, which resulted in high staff turnover and a high degree of change in administrative practices and staff mobilization, including transfers of heads of health centres. Three health centres reported financial constraints, high staff mobility and multiple smaller projects that made its management reluctant to embark on a new project, resulting in a decline to continue with the trial. The SMART2D steering committee took the decision in 2017 to halt further negotiations and continue with the two committed health centres.

Thus, the adaptive implementation trial originally proposed for the Swedish setting was converted to a feasibility trial [29]. Based on Bowen et al [29], the need to establish additional community partnerships to support prevention efforts beyond the facility-based primary care set-up in Sweden; the limited published studies and the need to understand the unique considerations for the implementation of complex health interventions studies in superdiverse neighbourhoods, fulfilled indications for performing a feasibility study and supported the rationale for this change in the Swedish context [33].

## Data Analysis

We used the intention-to-treat (ITT) analysis strategy where all participants were analysed as randomized, and those lost to follow-up were assumed to not have achieved the outcome of interest had they been followed to completion. The outcomes of interest as stated in the trial protocol [31], were:

Primary outcomes of interest

a. Among participants with T2D, the primary outcome was glycaemic control defined as having an HbA1c < 48 mmol/mol [48,49] at endline.
b. Among participants at high risk for T2D, the primary outcome was reduction in HbA1c between baseline and endline. We hypothesized that participants at high risk for T2D in the integrated care arm would on average have a higher reduction in their HbA1c level by the end of follow-up compared to high-risk participants in the facility-only arm.

Secondary outcomes of interest:

a. Retention into care among participants with T2D and those at high risk of T2D, defined as having an HbA1c measurement at both baseline and at endline.
b. Incidence of diabetes among participants at high risk for T2D, defined as having an endline, HbA1c reading of >= 48 mmol/mol.

A difference in differences analysis was conducted, i.e., a difference between the study arms, of differences between month 12 and baseline measurements. To compare the difference in percentage of participants with the outcomes of interest, we used the z-test statistic, and for multi-category variables the chi-square test, or Fishers’ Exact test where small expected-frequencies occurred in the tabulations. To compare continuous outcomes, we calculated the mean and the respective standard deviation (SD), or the median values of these outcomes, and used the Kruskal Wallis non-parametric test to compare differences in changes between the study arms. We report probability values (p-values) of these tests.

For adjusted analyses, and to minimize the effect of within cluster correlations (unit of analysis was individual participants), we used mixed effects multi-level modelling to adjust for health facility and individual levels. The data had a two-level hierarchical structure, i.e. individual participants (level 1) nested within health facilities (level 2). Two different types of models were used for adjusted analyses based on the type of outcome. For outcomes indicating the percentage with glycaemic control, retention to care, and incident diabetes; modified Poisson regression mixed effects modelling was applied. The cluster indicator was included in the model as the random effect, and the other variables, including the study arm and participants characteristics (age, sex, level of schooling completed, marital status, baseline BMI, baseline tobacco use status, baseline alcohol use status, baseline physical activity levels, baseline HbA1c, baseline dietary score, and baseline hypertension status) as fixed effects. Modified Poisson regression was chosen for these outcomes over logistic regression to avoid known underestimation of the standard errors for the estimated risk ratios when the outcome prevalence exceeds 10% [50,51]. From this modelling, we report the estimated incident rate ratios (IRRs) and their respective 95% confidence intervals (CI). For the outcome of reduction in HbA1c among participants at high risk for T2D, a difference-in-differences analysis was conducted of differences between month 12 and baseline measurements; and we applied the linear mixed-effects model, with the cluster variable as the random effect, and participant characteristics (see above) as fixed effects. For this modelling, we report the estimated reduction in HbA1c among participants in the integrated care arm, per unit reduction in HbA1c among participants in the facility only arm. In all adjusted analyses modelling, step-by-step backward elimination of variables was used, removing variables with the highest p-values one at time, and finally retaining only variables with p-values < 0.05 in the final model.

## RESULTS

### Enrolment and follow-up

A total of 142 participants were enrolled, 58 participants with T2D and 84 at high risk of T2D. Overall completion rates were 83% for participants with T2D 74% for participants at high risk of T2D. Table 4 summarizes the enrolment and follow-up completion rates and the distribution of participants enrolled by cluster. Among participants with T2D, the rate of completion was not significantly different between the facility-only arm (86.7%), and integrated care arm (78.6%) (p=0.415). Similarly, among participants at high-risk for T2D, we observed no significant difference in the completion rates between the study arms at 71.4% and 76.2%, respectively (p=0.620).

**Table 4.**
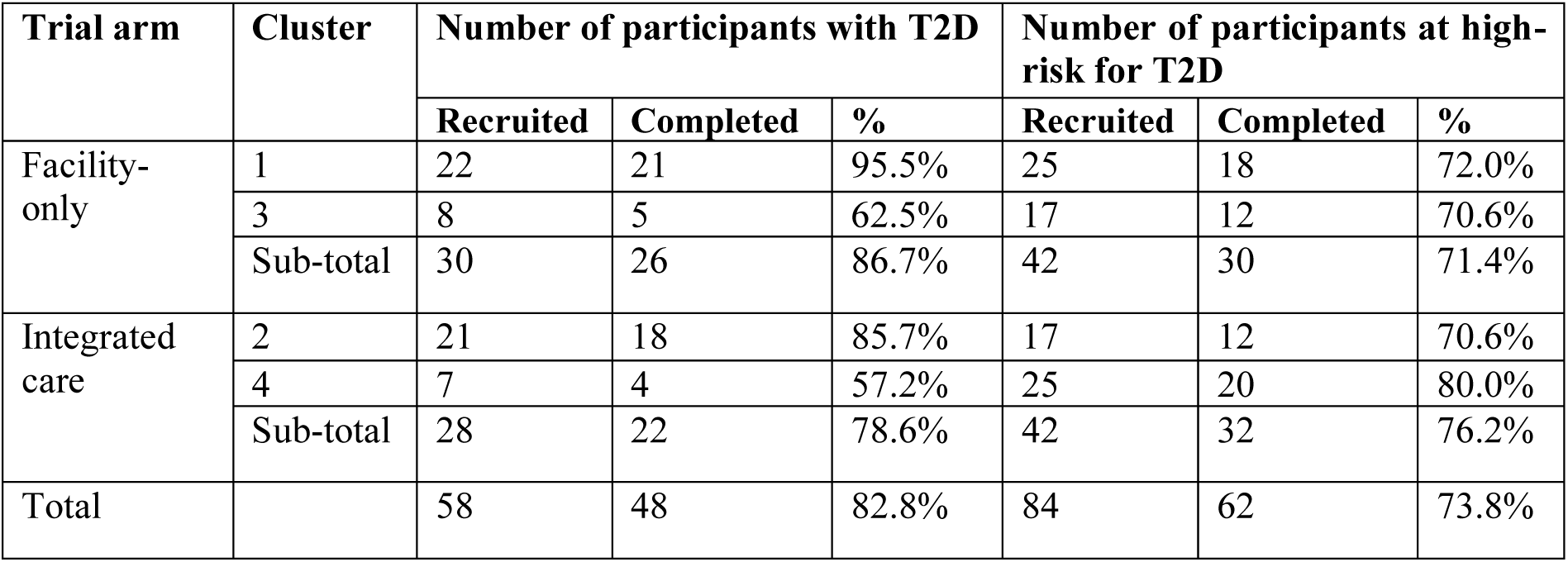
Number of participants by study arm and cluster, and study completion rates

### Baseline characteristics of participants

Table 5 describes the participants’ characteristics at baseline in terms of demographic, behavioural and biochemical variables, by study arm. Gender differences were observed in recruitment rates with a significantly higher proportion of women in the integrated care arm (81%) compared to the facility-only arm (60%) among those at high-risk for T2D. Participation rates for men and women were similar among participants with T2D in both arms. Participants were significantly younger in the integrated care arm across both T2D and high-risk categories. Behavioural and biochemical characteristics were largely comparable at baseline across both arms with some differences between T2D and high-risk categories within each arm. Overall, we observed no significant differences in the distribution of participant characteristics by study arm, both among participants with T2D and those at high risk of T2D.

**Table 5.** Baseline characteristics of participants

| Characteristic | Participants with T2D |  | Participants at high-risk for T2D |  |
| --- | --- | --- | --- | --- |
|  | Facility-only<sup>a</sup><br>(n=30) | Integrated care<sup>a</sup><br>(n=28) | Facility-only<sup>a</sup><br>(n=42) | Integrated care<sup>a</sup><br>(n=42) |
| Demographic characteristics |  |  |  |  |
| Sex |  |  |  |  |
| Females | 11 (36.7%) | 14 (50.0%) | 25 (59.5%) | 34 (81.0%) |
| Males | 19 (63.3%) | 14 (50.0%) | 17 (40.5%) | 8 (19.1%) |
|  | *p* = 0.306 |  | *p* = 0.032 |  |
| Age in years |  |  |  |  |
| 30-45 | 1 (3.3%) | 6 (21.4%) | 7 (16.7%) | 18 (42.9%) |
| 46-60 | 13 (43.3%) | 12 (42.9) | 16 (38.1%) | 18 (42.9%) |
| 60-75 | 16 (53.3%) | 10 (35.7%) | 19 (45.2%) | 6 (14.3%) |
|  | *p* = 0.085 |  | *p* = 0.003 |  |
| Mean ± SD | 60.1 ± 8.6 | 54.9 ± 11.3 | 56.5 ± 10.9 | 50.0 ± 12.5 |
|  | *p* = 0.049 |  | *p* = 0.013 |  |
| Place of birth according to UN regions<sup>b</sup> |  |  |  |  |
| Sweden | 12 (40.0%) | 11 (39.3%) | 11 (26.2%) | 10 (23.8%) |
| Other European countries | 8 (26.7%) | 2 (7.1%) | 5 (11.9%) | 0 (0.0%) |
| Africa | 4 (13.3%) | 6 (21.4%) | 10 (23.8%) | 17 (40.5%) |
| Americas | 0 (0.0%) | 1 (3.6%) | 2 (4.8%) | 3 (7.1%) |
| Asia | 6 (20.0%) | 8 (28.6%) | 14 (33.3%) | 12 (28.6%) |
|  | *p* = 0.249 |  | *p* = 0.123 |  |
| Education |  |  |  |  |
| Grade 0 - 12 | 10 (33.3%) | 11 (39.3%) | 15 (35.7%) | 16 (38.1%) |
|  | Facility-only<sup>a</sup><br>(n=30) | Integrated care<sup>a</sup><br>(n=28) | Facility-only<sup>a</sup><br>(n=42) | Integrated care<sup>a</sup><br>(n=42) |
| >12 and up to vocational training (before university) | 11 (36.7%) | 10 (35.7%) | 10 (23.8%) | 15 (35.7%) |
| University or higher | 9 (30.0%) | 7 (25.0%) | 17 (40.5%) | 11 (26.2%) |
|  | p = 0.871 |  | p = 0.314 |  |
| Marital status |  |  |  |  |
| Never married | 4 (14.3%) | 6 (21.1%) | 4 (9.5%) | 4 (9.5%) |
| Married/cohabiting | 20 (71.4%) | 17 (60.7%) | 16 (38.1%) | 21 (50.0%) |
| Separated/divorced | 4 (14.3%) | 5 (17.9%) | 18 (42.9%) | 14 (33.3%) |
| Widowed/ Other | 2 (6.7%) | - | 4 (9,5%) | 3 (7.1%) |
|  | p = 0.566 |  | p = 0.723 |  |
| Employment |  |  |  |  |
| Currently employed | 15 (50.0%) | 18 (64.3%) | 21 (50.0%) | 21 (50.0%) |
| Currently unemployed | 3 (10.0%) | 2 (7.1%) | 5 (11.9%) | 6 (14.3%) |
| Retired | 9 (30.0%) | 5 (17.9%) | 9 (21.4%) | 7 (16.7%) |
| Other | 3 (10.0%) | 3 (10.7%) | 7 (16.7%) | 8 (19.1%) |
|  | p = 0.669 |  | p = 0.939 |  |
| Behavioural & biochemical characteristics |  |  |  |  |
| Body Mass Index |  |  |  |  |
| <25.0 | 5 (16.7%) | 3 (10.7%) | 3 (7.1%) | 5 (11.9%) |
| 25.0-29.0 | 13 (43.3%) | 15 (53.6%) | 15 (35.7%) | 14 (33.3%) |
| ≥30.0 | 12 (40.0%) | 10 (35.7%) | 24 (57.1%) | 23 (54.8%) |
|  | p = 0.685 |  | p = 0.757 |  |
| Mean ± SD | 30.5 (±6.4) | 28.9 (±4.0) | 30.7 (±5.4) | 30.6 (±7.3) |
|  | p = 0.248 |  | p = 0.925 |  |
| Physical Activity % <sup>c</sup> |  |  |  |  |
| Yes | 16 (53.3%) | 17 (60.7%) | 20 (47.6%) | 24 (57.1%) |
| No | 14 (46.7%) | 11 (39.3%) | 22 (52.4%) | 18 (42.9%) |
|  | p = 0.571 |  | p = 0.382 |  |
| Mean ± SD | - | - | - | - |
| Current Tobacco Use % |  |  |  |  |
| Yes | 8 (26.7%) | 6 (21.4%) | 4 (9.5%) | 9 (21.4%) |
| No | 22 (73.3%) | 22 (78.6%) | 38 (90.5%) | 33 (78.6%) |
|  | p = 0.641 |  | p = 0.227 |  |
| Alcohol use % |  |  |  |  |
| 1-7 days/week | 12 (40.0%) | 14 (50.0%) | 17 (40.5%) | 9 (21.4%) |
| < 4 days/month | 6 (20.0%) | 4 (14.3%) | 3 (7.1%) | 5 (11.9%) |
| Never | 12 (40.0%) | 10 (37.7%) | 22 (52.4%) | 28 (66.7%) |
|  | p = 0.716 |  | p = 0.173 |  |
| HbA1c (mmol/mol) in Mean ± SD |  |  |  |  |
| HbA1c | 6.70 (±1.05) | 6.96 (±1.26) | 5.8 (±0.47) | 5.8 (±0.55) |
|  | p = 0.391 |  | p = 0.876 |  |
|  | Facility-only<sup>a</sup><br>(n=30) | Integrated care<sup>a</sup><br>(n=28) | Facility-only<sup>a</sup><br>(n=42) | Integrated care<sup>a</sup><br>(n=42) |
| **Hypertensive (systolic ≥ 140 mmHg, and/ or diastolic ≥ 90, and/ or on antihypertensive medication) %** |  |  |  |  |
| Normal | 6 (20.0%) | 9 (32.1%) | 8 (19.1%) | 10 (23.8%) |
| Hypertensive | 24 (80.0%) | 19 (67.9%) | 34 (81.0%) | 32 (76.2%) |
| | $p = 0.291$ | | $p = 0.595$ | |
| **Waist Circumference** |  |  |  |  |
| < 91 cm | 6 (20.0%) | 8 (28.6%) | 5 (11.9%) | 11 (26.2%) |
| ≥ 91 cm | 24 (80.0%) | 20 (71.4) | 37 (88.1%) | 31 (73.8%) |
| | $p = 0.446$ | | $p = 0.095$ | |

<sup>a</sup> Column percentages used. <sup>b</sup> Place of birth according to UN regions: **Sweden**; **Other European countries** (Eastern, Western, Northern, Southern Europe sub-regions, excluding Sweden); **Africa** (Northern and Sub-Saharan Africa); **Americas** (mainly Latin America and the Caribbean sub-region); and **Asia** (Predominantly Southern and Western Asian sub-regions). <sup>c</sup> Physical Activity %: Equivalent to at least 150 mins of moderate PA, or 75 mins of vigorous PA / week

### Effectiveness of the SMART2D intervention on primary outcomes

#### Glycaemic control among participants with T2D

Among 28 participants with T2D enrolled in the integrated care arm, 16 (57.1%) achieved glycaemic control by endline as compared to 19 out of 30 participants (63.3%) in the facility only arm (p=0.630). Adjusted analysis revealed a significantly lower rate of glycaemic control in the integrated care arm as compared to the facility only arm, with an adjusted IRR of 0.92, and a 95% CI= [0.89 – 0.95] (Table 6).

**Table 6.**
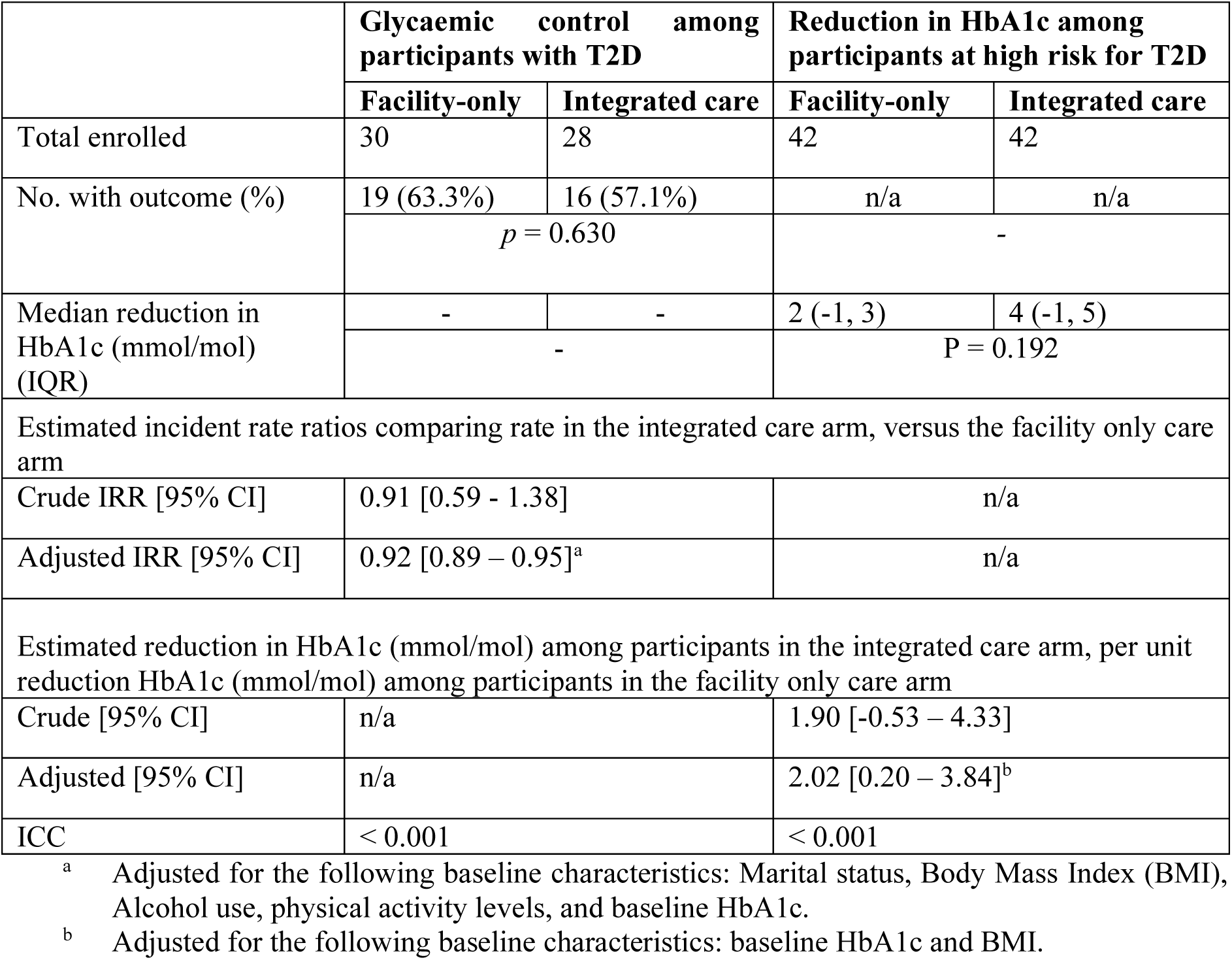
Glycaemic control among participants with T2D, and reduction in HbA1c among participants at high risk of T2D, in facility-only and integrated care arms

#### Reduction in HbA1c among participants at high risk of T2D

Among participants at high risk for T2D, the median reduction in HbA1c levels between baseline and endline was 4 mmol/mol in the integrated care arm, compared to a median reduction of 2 mmol/mol in the facility only arm (p-value= 0.192). Adjusted analysis showed a reduction of 2.02 mmol/mol among participants in the integrated care arm, per unit reduction in HbA1c among participants in the facility only care arm, which was marginally significant with a 95% CI of [0.20 – 3.84] (Table 6).

### Effectiveness of the SMART2D intervention on secondary outcomes

#### Retention into care

Among 28 participants with T2D in the integrated care arm, 22 (78.6%) were retained into care by endline as compared to 26 out of 30 participants (86.7%) in the facility only arm (p=0.499). Adjusted analysis showed a significantly lower rate of retention into care in the integrated care arm as compared to the facility only arm, with an adjusted IRR =0.91 [0.91 – 0.92]. Among participants at high risk for T2D, of the 42 participants in the integrated care arm, 32 (76.2%) were retained into care by endline as compared to 30 out of 42 participants (71.4%) in the facility only arm (p=0.620). In the adjusted analysis, a significantly higher rate of retention into care was observed in the integrated care arm as compared to the facility only arm with an adjusted IRR = 1.22 [1.15 – 1.29] (Table 7).

**Table 7.** Retention to care in facility only and integrated care arms by baseline T2D status

|  | Retention into care among participants with T2D |  | Retention into care among participants at high risk of T2D |  |
| --- | --- | --- | --- | --- |
|  | Facility-only | Integrated care | Facility-only | Integrated care |
| Total enrolled | 30 | 28 | 42 | 42 |
| No. retained to care (%) | 26 (86.7%) | 22 (78.6%) | 30 (71.4%) | 32 (76.2%) |
| | $p = 0.499$ | | $p = 0.620$ | |
| Crude IRR [95% CI] | 0.91 [0.71 - 1.15] |  | 1.07 [0.83 - 1.38] |  |
| Adjusted IRR [95% CI] | 0.91 [0.91 - 0.92]<sup>a</sup> |  | 1.22 [1.15 - 1.29]<sup>b</sup> |  |
| ICC | < 0.001 |  | < 0.001 |  |

<sup>a</sup> Adjusted for the following baseline characteristics: tobacco use
<sup>b</sup> Adjusted for the following baseline characteristics: age group, marital status, tobacco use, alcohol use, and Hypertension status

#### Incidence of diabetes among participants at high risk for T2D

Of the 42 participants at high risk for T2D in the integrated care arm, we did not observe any incident case of diabetes by endline, whereas among the 42 participants at high risk of T2D in the facility-only care arm, we observed two incident cases. Because of absence of cases in one of the study arms, it was not possible to conduct adjusted analysis for this secondary study outcome.

## DISCUSSION

The main findings of the study are: 1) The change to a feasibility trial to study proof-of-implementation of the SMART2D intervention in superdiverse and socioeconomically disadvantaged urban areas shed light on contextual challenges relevant for implementation of prevention interventions in hard-to-reach urban neighbourhoods; 2) For participants at high-risk of T2D, the integrated care arm was found to be more effective in improving retention to care and reduction in HbA1c, as compared to the facility-only arm; 3) The reverse was true for participants with T2D, where the facility only arm was effective in achieving glycaemic control and better retention into care as compared to the integrated care arm.

### Feasibility trials for improving real-world relevance

The present trial tested the feasibility of a SMART2D real-life complex health intervention in superdiverse neighbourhoods to assess its potential to proceed to full implementation. Other studies that are a part of this evaluation assessed the reach of community-based screening in comparison to facility-based opportunistic screening; the acceptability of the tele-health coaching component; and, its fidelity, including the quality and consistency of interactions between participants and facilitators [41,43,43]. Apart from the findings of this study, we also identified that the superdiversity in the target population manifested as patterns of multiple needs, which must be better addressed in the content and delivery of the tele-health coaching and community outreach components. While the overall structure of the tele-health coaching was adequate, it lacked flexibility to support the wide spectrum of needs contained in the target population. We also identified differences in how facilitators supported goal setting by participants, which indicate a need for focused training on goal setting and related components of the intervention, in line with the findings of a systematic review by Pinto et al [52]. which found inconsistencies in goal setting, including poor standardization in terms of component description and reporting for chronic disease prevention and management interventions in sub-Saharan Africa. Facilitators in that study also reported specific practical challenges, including difficulties in adapting standard goal-setting frameworks (e.g., Specific, Measurable, Achievable, Relevant and Time-bound - SMART), and navigating group-based delivery versus one-to-one dialogue with individual participants or couples [52]. This study hence provides a case-in-point for the conduct of more feasibility trials in real-life contexts in the framing of implementation research.

It is important to recognise and emphasise that assessing the feasibility of implementing a complex intervention from different perspectives as described elsewhere [41–43] is as important as evaluating its effectiveness. The real-world implementation of evidence-based practices often lack elements that feasibility trials include and can in turn help to strengthen the process of implementation [53,54]. A community-based intervention to prevent T2D in high-risk adults in England for example, followed its participants over the course of 12-months to assess the feasibility, accessibility and outcomes of the study. Apart from the high rates of retention and positive outcomes in physical activity levels, weight loss, reduction in waist circumference and FINDRISC score; the study showed encouraging indications of the feasibility and implementation potential of the intervention. In addition, they identified poor participation among ethnic minorities, despite their initial objective of recruiting participants in socioeconomically deprived areas [55].

### Primary care and community outreach for T2D prevention and management

The feasibility trial targeted participants with diagnosed T2D and participants at high risk of T2D. In the Swedish healthcare setting, those with diagnosed T2D would have regular follow-up for medication and testing through their primary care as well as prescribed medication and home-testing equipment for HbA1c or blood glucose through subsidised pharmacy services [38]. Those with high risk of T2D on the other hand had no access to regular subsidized care or support since they did not have a diagnosis code. Within the high-risk category however, those who fulfilled clinical parameters for pre-diabetes had access to primary care and lifestyle support. In theory, all residents had the right to access primary care and lifestyle support even without a diagnosis, but in reality these numbers are very low and waiting lists are long [56].

We found that the facility only care arm showed better retention to care and glycaemic control for participants with T2D, compared to the integrated care arm. Among high-risk participants, a greater proportion in the integrated care arm showed HbA1c reduction of at least 3 mmol/mol. These findings underscore the role of community outreach including tailored personal coaching in T2D prevention among those at high risk and that of a well-functioning primary care in T2D management for those with a diagnosis. It is well documented that healthcare services, including primary care, do not have the financial or human resources to provide adequate preventative services for chronic health conditions [43].The cost of providing these services is globally prohibitive, irrespective of country income status [57]. Additionally, access to available services has been found to be disproportionately low among hard-to reach population groups, including in Sweden, thereby increasing their vulnerability [58]. Previous studies indicate that interventions implemented in hard-to-reach communities can however be successful and provide positive results for diabetes prevention [59]. This feasibility trial shows that preventative services tailored to community and individual needs through personal coaching and community outreach can provide a viable option for community-dwelling adults in socio-economically disadvantaged neighbourhoods. Multi-stakeholder engagement and utilising existing social capital in the form of community resources and engaged civil society organisations can potentially improve the cost-effectiveness of such interventions at scale [60].

### Socio-demographic diversity in target population

In this feasibility trial, more women, younger participants, and non-European participants were included in the integrated care arm compared to the facility only care arm. Since most participants (excluding those with T2D) were recruited through open community screening, this difference in distribution reflects the high prevalence or risk of T2D in socio-economically disadvantaged neighbourhoods with a high proportion of non-European immigrants, who may also have a genetic predisposition to T2D [61–63] or be affected by epigenetic imprinting [64] linked to increased T2D risk. Intervention development and implementation become more complex in the context of diverse populations[65]. The distribution of age, gender and place of birth in this study reflects our earlier studies on screening within the same project, where Timm at al. reported that women were significantly over-represented in both facility and community-based screening for T2D; while younger participants and those with a non-European background were reached more through community-based screening [43].

Chen et al found that interventions for T2D prevention focusing on lifestyle are broadly effective across ethnic groups, though the magnitude of benefit on glycemic and anthropometric outcomes varies by ethnicity, with European, South Asian, East/Southeast Asian, Middle Eastern, Latin American, and African groups showing differing responses [66]. A related publication on analysis of intervention design features found that a higher number of intervention sessions was significantly associated with greater reductions in both diabetes incidence and weight, while other characteristics such as who delivered the intervention and its delivery format, showed no significant effect, regardless of ethnic group [67]. Together, the two papers suggest that while ethnicity shapes the degree of physiological benefit from lifestyle interventions, intervention intensity (session frequency) was the design factor most consistently linked to better outcomes across ethnic groups [66,67].

### Methodological considerations

As outlined in the methods, the feasibility trial deviated significantly from the original SMART2D protocol which proposed a full trial in three settings (Uganda, South Africa, Sweden) due to significant administrative and contextual challenges in the Swedish site. While the feasibility study maintained fidelity to the core outcomes, it placed greater emphasis on the process of implementation through qualitative and mixed-methods evaluation. The feasibility trial also saw key modifications in study design, including a shift from a peer-group intervention to tele-health coaching, while maintaining the original strength-based approach [29]. The eligibility criteria were broadened to include individuals at high risk for diabetes, as many participants did not return for confirmatory diagnoses, leading to delays and loss to follow-up. Field implementation requires that processes are simplified and streamlined while maintaining the necessary conditions. Additionally, the intervention duration was reduced from one year to three months to align with feasibility study constraints, necessitating adjustments in sample size, outcome measures, and intervention completion criteria. These deviations highlight the challenges of conducting community-based diabetes prevention interventions in complex and evolving healthcare environments.

### Future implications

In this trial, we were able to test the limited effectiveness of a complex health intervention in superdiverse neighbourhoods where ethnic diversity is only one aspect of the diversity contained within the population. Language as a critical factor was addressed through special language facilitators for the tele-health coaching. However, the availability of all tools in multiple languages and further improving the scope of multi-language interactions overall is crucial in any subsequent iteration of the intervention. Across this study and related studies, we identified critical gaps in implementation processes [41–43]. We also identified the need to stratify the intervention content and delivery to address the spectrum of needs in the target population. The results of the feasibility trial together with the identified gaps have resulted in an ongoing project, PREVENT, where the focus is on healthy eating and active living for the prevention of cardiometabolic diseases for individuals at high risk [68]. Beyond this study, the findings highlight the importance of feasibility trials to strengthen implementation processes and outcomes. There is an increasing need to address common risk factors for chronic health conditions through common frameworks independent of available therapeutic modalities. An intervention such as tested here provides a flexible and modular framework that could be adapted to address multiple risk factors. Additionally, funders should consider promoting interim studies such as these, which are critical to address prevention gaps through innovative solutions that straddle the bounds of formal healthcare settings.

## CONCLUSIONS

The SMART2D feasibility trial offers proof-of implementation of a real-world complex health intervention, specifically for the prevention of T2D and potentially other chronic health conditions in hard-to-reach neighbourhoods. The role of interim feasibility trials along the effectiveness-implementation-scale-up pipeline remains valuable, as it sheds light on the interactions between the intervention and contextual elements within the target population to address prevention gaps from individual and societal perspectives.

## Supporting information

CONSORT 2010 extension checklist for pilot and feasibility trials

## Data Availability

The data supporting the findings of this study are available from the corresponding author upon reasonable request.

## Acknowledgements

SMART2D contributors (in alphabetical order of last names): Jhon Alvarez Ahlgren, Juliet Aweko, Aravinda Berggreen Clausen, Jeroen De Man, Ida Karlsson, Karima Lundin, Stefan Swartling Peterson, Göran Tomson, Claes-Göran Östenson We would like to thank the participants of the feasibility trial; local stakeholders who participated in the trial planning workshops; the personnel from the cooperating primary health care centres and the field workers who conducted the T2D screening; and the citizen service offices and civil society organisations who helped to mobilize the community during screening. We would also like to thank Jing Qi for editorial support in the current version of the manuscript and Mathilde Sengoelge and Johanna Stjärnfeldt for editorial support in previous versions.

## Author contributions (based on CReDiT Classification)

Meena Daivadanam: Conceptualisation, Funding Acquisition, Development of Methodology, Investigation, Project Administration, Supervision, Validation, Writing – Original draft preparation, review and editing

Kristi Sidney Annerstedt: Data Curation, Data Collection (including quality checks), Data management, Supervision of data collection, Formal analysis, Writing – review and editing

Helle Mölsted Alvesson: Conceptualisation, Development of Methodology, Supervision, Writing – review and editing

Linda Timm: Data Collection, Field implementation of project, Field supervision and quality checks, Writing – review and editing

Carl Johan Sundberg: Conceptualisation, Funding Acquisition, Development of Methodology, Writing – review and editing

Clas-Göran Östenson: Conceptualisation, Funding Acquisition, Development of Methodology, Writing – review and editing

Peter Delobelle: Conceptualisation, Funding Acquisition, Development of Methodology, Writing – review and editing

Josefien Van Olmen: Conceptualisation, Funding Acquisition, Development of Methodology, Writing – review and editing

Pilvikki Absetz: Conceptualisation, Funding Acquisition, Development of Methodology, Writing – review and editing

David Guwatudde: Conceptualisation, Funding Acquisition, Data Curation, Development of Methodology, Formal Analysis, Supervision, Validation, Visualisation, Writing – Original draft preparation, review and editing

## Ethics and consent

All procedures performed as part of the SMART2D project involving human participants were in accordance with the ethical standards of the institutional and/or national research committees involved and with the 1964 Helsinki declaration and its later amendments or comparable ethical standards. The SMART2D study was approved by the ethics committee in each of the respective countries, which for Sweden was the Regional Ethics Review Board in Stockholm (Ref 2015/712-31/1). Informed consent was obtained from all individual participants included in the study.

## Funding information

This study is part of the SMART2D project funded by the European Commission’s Horizon 2020 Health Coordination Activities (Grant Agreement No 643692) under call “HCO-05-2014: Global Alliance for Chronic Diseases: Prevention and treatment of type 2 diabetes”. The SMART2D consortium included the following six partner institutions: Karolinska Institutet, Sweden; Institute of Tropical Medicine, Belgium; Collaborative Care Systems, Finland; Makerere University, School of Public Health, Uganda; University of Western Cape, School of Public Health, South Africa; and Uppsala University, Sweden. The content of this article is solely the responsibility of the authors and does not reflect the views of the EU. The funders had no role in the study design, data collection and analysis, decision to publish or preparation of the manuscript.

Paper context:

Data availability statement: ORCID identifiers: Preprints:

Supplemental online material:

